# High incidence of pulmonary thromboembolism in hospitalized SARS-CoV-2 infected patients

**DOI:** 10.1101/2021.03.23.21253258

**Authors:** D El-Qutob, L Alvarez, P García-Sidro, M Robustillo, I Barreda, M Nieto, M Pin, FJ Carrera-Hueso

**Affiliations:** Unit of Allergy, Universitary Hospital of La Plana in Vila-Real, Spain; Service of Pharmacy, Universitary Hospital of La Plana in Vila-Real, Spain; Doctoral Program of Pharmacy, Universitary of Granada, Spain; Section of Pneumology, Universitary Hospital of La Plana in Vila-Real, Spain; Unit of Rheumatology, Universitary Hospital of La Plana in Vila-Real, Spain; Section of Neurophysiology, Universitary Hospital of La Plana in Vila-Real, Spain; Unit of Nephrology, Universitary Hospital of La Plana in Vila-Real, Spain

**Keywords:** coronavirus, COVID-19, SARS-CoV-2, coagulation, thrombosis, thromboembolism

## Abstract

**Introduction:** SARS-CoV-2 infected patients present thrombotic complications caused by direct endothelial cells injury of the microvessels. Pulmonary thromboembolism (PE) has been reported by Computed Tomography pulmonary angiogram (CTPA) in patients with COVID-19 pneumonia with high D-dimer levels.

**Objectives:** We present the characteristics of SARS-CoV-2 infected patients diagnosed of PE by CTPA in our hospital. We also present the comparison of these findings with non-infected patients with PE data.

**Methods:** Patients 18 years of age or older with SARS-CoV2 virus infection, and patients with suspected infection at beginning of admission but with negative PCR, were studied with CTPA for suspicion of VTE, during their hospitalization.

**Results:** During the study period, 52 CTPA were performed in our hospital, sixteen in SARS-CoV-2 infected patients. No significant differences in age (p=0.43) and sex (p=0.31) were found between the two groups, infected and non-infected patients. In the infected group, the patients who had PE had a much lower median age (47.8 years) than those without PE (73.3 years). No differences between infected and non-infected patients were detected in the diagnosis of PE with CTPA, 28.6% versus 27.8% (p=1.00). Overall patient mortality was 1.9%; one patient died (6.3%) in the infected group, and none in the non-infected group (p=0.31).

**Conclusion:** A considerable incidence of PE diagnosed by CTPA in SARS-CoV-2 infected patients has been observed, despite thrombo-prophylaxis.

## Introduction

The outbreak of the COVID-19 pandemic, caused by the novel coronavirus Severe Acute Respiratory Syndrome-CoronaVirus-2 (SARS-CoV-2), in the city of Wuhan, Hubei province of China, was declared by the World Health Organization (WHO) on March 21st. Thrombotic complications are an important issue in patients infected with COVID-19. Pulmonary thromboembolism (PE) has been reported by computed tomography pulmonary angiogram (CTPA) in patients with COVID-19 pneumonia with high D-dimer levels^1^ and in COVID-19 patient with normal D-dimer level, without strong predisposing risk factors for venous thrombo-embolism (VTE)^2^.

SARS-CoV-2 binds Angiotensin-converting enzyme 2 receptors through the viral surface spike protein and gain entry to cells. This protein, higher in individuals with cardiovascular disease, is linked by plasmin, and potentially making them more susceptible to worse outcomes^3^. This pattern of prothrombotic coagulopathy is different from what noticed in sepsis, where thrombocyte count is usually decreased, and of disseminated intravascular coagulation, where the exhausted coagulation system shows a prolongation of the prothrombin time and aPTT (activated Partial Thromboplastin Time), and a haemorrhagic tendency^4^. SARS-CoV-2 infection is suspected of producing deregulation of the coagulation system, with formation of intra-alveolar or systemic fibrin clots, associated with severe respiratory disease^5^. SARS-CoV-2 produces direct endothelial cells injury of the microvessels and then releases these damaged endothelial cells into the circulation^6^. PE has been reported in patients suffering from SARS-CoV-2 infection (2). Moreover, microthrombosis of small pulmonary vessels has been observed in the post-mortem inspection of lungs^7^. Immobilization due to prolonged intensive care unit (ICU) admission in severely infected patients as well as a hypercoagulable state caused by SARS-CoV-2, may be relevant factors.

Consequently, PE may be considered in SARS-CoV-2 infected patients with abrupt onset of oxygenation desaturation, respiratory distress, and decreased blood pressure^8^. This possibility might be suspected also by elevated D-dimer values, even though D-dimer is a non-specific acute phase reactant. However, other authors argued that there is no evidence for use of biomarkers such as D-dimer to guide intensification of anticoagulant dosing despite it being a marker of poor prognosis^9^.

In a recent study investigating the prognostic factors of 28-day mortality of severely SARS-CoV-2 infected patients, the use of anticoagulant therapy for at least seven days, resulted in lower mortality in patients with D-dimer over six fold the upper limit of normal (Low Molecular Weight Heparin (LMWH): 32.8% vs No-LMWH: 52.4%, P=0.017), but without overall benefit for patients on LMWH^7^. A recent study examined two groups of patients: those with COVID-19 and those without COVID-19. The COVID-19 group showed lower mortality rates with heparin administration (LMWH (40–60 mg enoxaparin/day) or unfractionated heparin (UFH) (10,000– 15,000 U/day)) than those without heparin. Interestingly, there was no difference in mortality in the COVID-19 negative patients with the use of heparin^10^. Our goal is to analyze the characteristics of SARS-CoV-2 infected patients diagnosed of PE by CTPA in our hospital.

## Methods

### Study population

Patients 18 years of age or older with SARS-CoV2 virus infection confirmed by diagnostic test of reverse transcription of polymerase chain reaction (RT-qPCR) in a sample of nasopharyngeal or sputum aspirate, and patients with suspected infection at beginning of admission but with negative PCR, who were studied with CTPA for suspicion of VTE, during their ward admission to an European Hospital from February 26th- to May 20^th^ of 2020.

Patients who denied consent for CTPA, and pregnant or breastfeeding women were excluded from the study. Patients who were re-admitted at the hospital for any reason were also excluded, and only the first episode of hospitalization was considered.Our routine protocol for patients with severe clinical features of COVID-19 infection was multidetector pulmonary CT angiography using 16 slice multi-detector CT (GE Healthcare, Milwaukee, WI) after intravenous injection of 60 ml iodinated contrast agent (Iohexol 350 mg/mL, GE Healthcare, Milwaukee, WI) at a flow rate of 3.5 mL/s, triggered on the main pulmonary artery.

Demographic data, comorbidities, clinical symptoms, laboratory results, radiological tests and treatment of each patient were evaluated. A data collection sheet was created in Microsoft Excel where all variables were automatically collected. Statistical analysis and processing of the data was carried out using the SPSS version 19 statistical package (IBM Corp. Released 2010. IBM SPSS Statistics for Windows, Version 19.0. Armonk, NY: IBM Corp.). Categorical variables are represented as absolute frequencies and percentages, and quantitative variables are expressed by median and Interquartile Range (IQR) calculated by Tukey’s Hinges method. The difference analysis used student’s T test for variables or the Mann-Whitney test if they have normal distribution or not respectively. To contrast the categorical variables, the test of chi-squared test (χ2) The statistical significance level adopted for all contrast tests was p<0.05.

### Results

During the study period, 52 CTPA were performed in our hospital, sixteen in SARS-CoV-2 infected patients with positive RT-qPCR test. The median age of all patients included in the study was 70.1 years (Interquartile range (IQR): 55.7 to 83.3) and 27 (51.9%) were male. No significant differences in age (p=0.43) and sex (p=0.31) were found between the two groups, infected and non-infected patients. However, in the infected group, the patients who had PE had a much lower median age (47.8 years) than those without PE (73.3 years). Table 1 shows the demographic and comorbidities of the patients. The most common comorbidities were hypertension (71.2%), hyperlipidaemia (48.1%), diabetes (34.6%) and obesity (36.5%). In the non-infected group, congestive heart failure (27.8%, p=0.02) and previous treatment with anticoagulants (22.2%, p=0.04) were more significantly frequent than in the infected group.

**Table 1.**
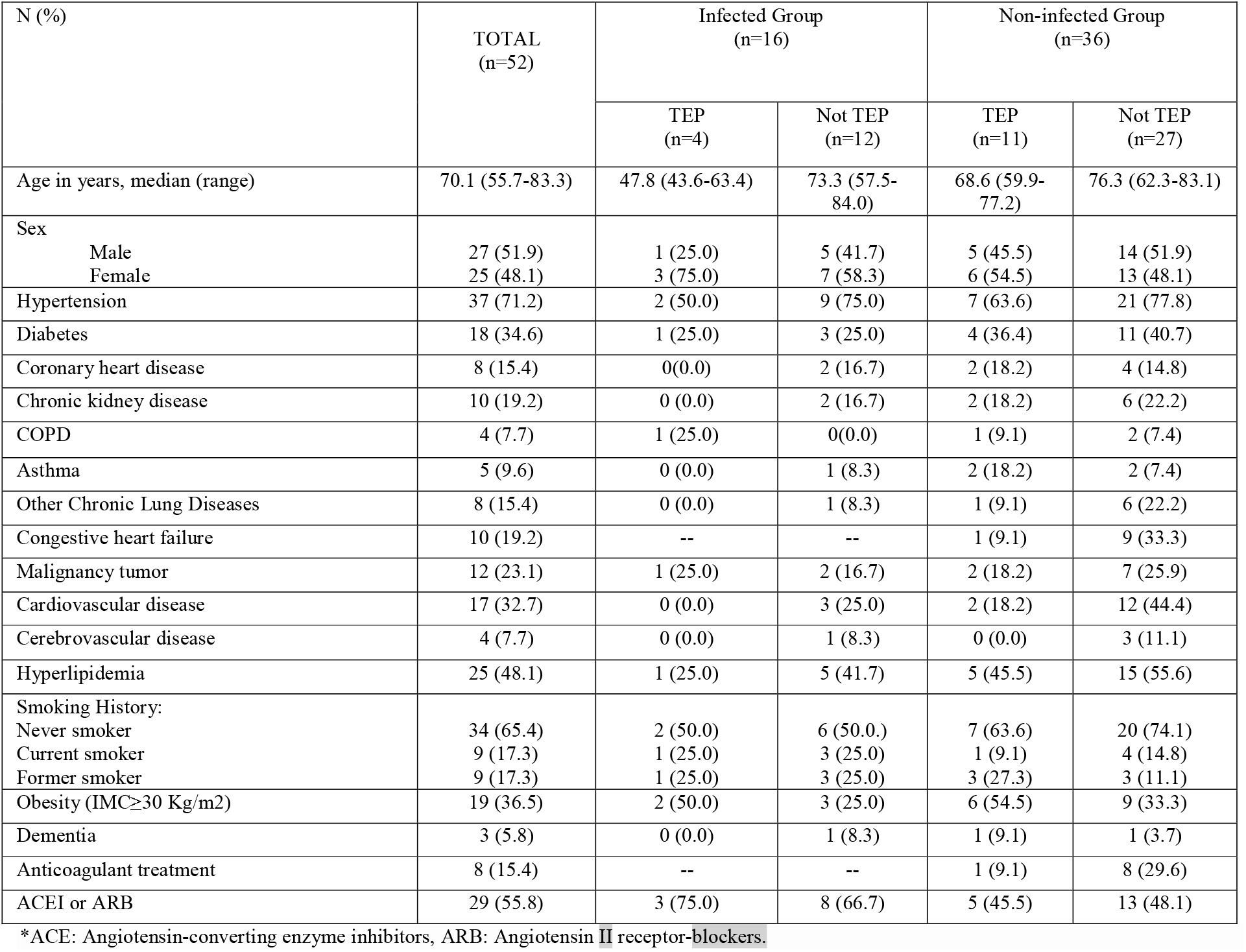
**Demographic characteristics and comorbidities of the patients stratified by negative or positive infection**

The median of total comorbidities was 2.5 (IQR:1.0 to 3.5) and 4.0 (IQR: 2.0 to 6.0) for infected and non-infected patients, respectively (p=0.07), and for all the patients was 3 (IQR: 2.0 to 5.5). According to the Charlson Comorbidity Index abbreviated, 53.8% of the patients (n=28) do not have comorbidities.

The most common symptoms were fever, dyspnea, malaise, and cough (see Table 2). Fever was more common in the infected group with statistically significant difference (75.0% versus 41.7%, p=0.03).

**Table 2.** Signs and symptoms of the patients on admission.

| N (%) | TOTAL<br>(n=52) | Infected Group<br>(n=16) |  | Non-infected Group<br>(n=36) |  |
| --- | --- | --- | --- | --- | --- |
|  |  | TEP<br>(n=4) | Not TEP<br>(n=12) | TEP<br>(n=11) | Not TEP<br>(n=27) |
| Fever | 27 (51.9) | 3 (75.0) | 9 (75.0) | 4 (36.4) | 11 (40.7) |
| Dyspnea | 28 (53.8) | 2 (50.0) | 6 (50.0) | 5 (45.5) | 15 (55.6) |
| Cough | 18 (34.6) | 2 (50.0) | 4 (33.3) | 4 (36.4) | 9 (33.3) |
| Expectoration | 5 (9.6) | 0 (0.0) | 1 (8.3) | 1 (9.1) | 4 (14.8) |
| Sore throat | 3 (5.8) | -- | -- | 0 (0.0) | 3 (11.1) |
| Myalgia | 9 (17.3) | 1 (25.0) | 2 (16.7) | 4 (36.4) | 3 (11.1) |
| Headache | 2 (3.8) | -- | -- | 1 (9.1) | 1 (3.7) |
| Dizziness | 6 (11.5) | 0 (0.0) | 2 (16.7) | 0 (0.0) | 4 (14.8) |
| Diarrhea | 7 (13.5) | 1 (25.0) | 2 (16.7) | 0 (0.0) | 4 (14.8) |
| Chest pain | 6 (11.5) | 0 (0.0) | 1 (8.3) | 1 (9.1) | 4 (14.8) |
| Malaise | 21 (40.4) | 2 (50.0) | 7 (58.3) | 3 (27.3) | 9 (33.3) |
| Anosmia | 2 (3.8) | 1 (25.0) | 1 (8.3) | -- | -- |
| Dysgeusia | 3 (5.8) | 1 (25.0) | 1 (8.3) | 0 (0.0) | 1 (3.7) |

Abnormal chest X-ray was observed in most patients at admission. Radiological findings are showed in Table 3. Chest CT scan was significantly different between infected and non-infected patients (p<0.01). The most common abnormality was ground-glass opacities in both groups, being more frequent in the infected group than in the non-infected (50% vs 11%). No differences between infected and non-infected patients were detected in the diagnosis of PE with CTPA, 28.6% versus 27.8% (p=1.00). Bilateral affectation (p=0.01), and the parenchymal lesion p<0.01) were significantly more frequent in the infected group (The median of hospitalization days before CTPA was performed, was 3.5 days (IQR: 1.0 to 9.0); being 2 (IQR: 0.5 to 6.0) and 9.5 (IQR: 3.5 to 13.0) for the non-infected group and infected group, respectively.

**Table 3.** Radiological findings.

| N (%) | TOTAL<br>(N=52) | Infected Group<br>(n=16) |  | Non-infected Group<br>(n=36) |  |
| --- | --- | --- | --- | --- | --- |
|  |  | TEP<br>(n=4) | Not TEP<br>(n=12) | TEP<br>(n=11) | Not TEP<br>(n=27) |
| Chest X-ray |  |  |  |  |  |
| Normal | 16 (30.8) | 2 (50.0) | 1 (8.3) | 7 (63.6) | 9 (33.4) |
| Opacities | 12 (23.1) | 1 (25.0) | 7 (58.3) | 1 (9.1) | 3 (11.1) |
| Interstitial pattern | 9 (17.3) | 1 (25.0) | 4 (33.3) | 1 (9.1) | 3 (11.1) |
| No COVID Injuries | 14 (26.9) | 0 (0.0) | 0 (0.0) | 2 (18.2) | 12 (44.4) |
| CTPA |  |  |  |  |  |
| Bilateralism | 32 (61.5) | 4 (100.0) | 10 (83.3) | 10 (90.9) | 9 (33.3) |
| Parenchymal injury | 14 (26.9) | 2 (50.0) | 7 (58.3) | 3 (27.3) | 2 (7.4) |
| Pleural effusion | 19 (36.5) | 0 (0.0) | 5 (41.7) | 0 (0.0) | 14 (51.9) |
| Pericardial effusion | 4 (7.7) | 0 (0.0) | 1 (8.3) | 0 (0.0) | 3 (11.1) |
| Metastasis | 3 (5.8) | 0 (0.0) | 1 (8.3) | 1 (9.1) | 1 (3.7) |

Table 4 shows the laboratory findings before CTPA. The most relevant laboratory findings were lymphopenia, lengthening of the prothrombin time, elevated LDH, D-dimer and ferritin. Figure 1 shows the levels of D-dimer categorized by groups and result of PE. Among the patients who had PE, statistically significant differences were found in the levels of total CK (p = 0.01) and troponin T (p = 0.02), being higher in the infected group compared to the non-infected.

**Table 4.** Laboratory results before CTPA.

| Laboratory results, median (IQR) | TOTAL (N=52) | Infected Group (n=16) |  | Non-infected Group (n=36) |  | Reference ranges |
| --- | --- | --- | --- | --- | --- | --- |
|  |  | TEP (n=4) | Not TEP (n=12) | TEP (n=11) | Not TEP (n=27) |  |
| White blood cell count, x 10<sup>9</sup>/L | 7.6 (5.9-9.8) | 7.7 (6.3-9.1) | 6.7 (4.3-8.4) | 9.0 (6.8-12.1) | 7.7 (5.5-9.9) | 4.8-11 |
| Hemoglobin, g/dL | 12.7 (11.1-14.3) | 11.8 (10.9-12.6) | 12.7 (12.6-14.7) | 12.9 (10.4-14.1) | 12.9 (11.0-14.2) | 12-18 |
| Neutrophil count, x 10<sup>9</sup>/L | 5.4 (3.3-8.0) | 5.3 (4.2-6.3) | 4.9 (1.7-5.4) | 7.2 (5.4-9.2) | 5.5 (3.1-8.0) | 1.9-8 |
| Lymphocyte count, x 10<sup>9</sup>/L | 1.30 (0.95-1.65) | 1.3 (1.0-1.6) | 1.4 (1.0-1.7) | 1.0 (0.7-1.8) | 1.3 (1.0-1.6) | 0.9-4.5 |
| Platelet count, x 10<sup>9</sup>/L | 204 (160-255) | 178 (152-204) | 263 (225-276) | 199 (174-312) | 198 (146-222) | 130-400 |
| D-dimer, µg/mL | 2323.5 (1083.0-7301.5) | 5824.5 (5120.0-6523.0) | 1573.0 (1041.0-5873.0) | 5078.5 (2258.0-12721.0) | 1731.0 (1007.5-2494.5) | 0-500 |
| Prothrombin time, sec | 12.5 (11.6-13.3) | 12.7 (12.0-13.4) | 11.7 (11.4-13.2) | 12.5 (11.2-12.9) | 13.0 (12.1-13.4) | 9-13 |
| Glucose, mg/dL | 118 (94-145) | 112 (109-115) | 102 (92-124) | 139 (118-181) | 105 (92-141) | 82-115 |
| Creatinine, mg/dL | 0.91 (0.76-1.10) | 1.40 (1.08-1.72) | 0.81 (0.68-0.87) | 0.90 (0.70-0.99) | 0.94 (0.83-1.15) | 0.7-1.2 |
| Total bilirubin, mg/dL | 0.40 (0.28-0.55) | 0.34 (0.20-0.48) | 0.47 (0.31-0.60) | 0.29 (0.22-0.45) | 0.41 (0.36-0.59) | 0.1-1.2 |
| Sodium, mmol/L | 140.2 (138.0-142.6) | 143.5 (142.2-144.8) | 142.0 (140.0-143.0) | 139.4 (138.5-142.6) | 139.7 (136.1-142.0) | 136-146 |
| Potassium, mmol/L | 4.52 (4.08-4.80) | 5.62 (4.61-6.62) | 4.40 (4.09-4.67) | 4.27 (4.01-4.47) | 4.57 (4.23-4.91) | 3.5-5.1 |
| Alanine aminotransferase (GPT), U/L | 21.5 (12.8-38.4) | 9.0 (8.3-9.6) | 36.3 (23.7-73.1) | 22.3 (17.2-28.3) | 18.7 (9.9-37.9) | 5-41 |
| Lactate dehydrogenase, U/L | 223 (199-262) | 273 (197-348) | 231 (204-260) | 223 (208-237) | 231 (164-276) | 135-225 |
| CK total, U/L | 61.0 (4.5-91.5) | 135.5 (102.0-169.0) | 61.0 (45.0-78.5) | 87 (33-99) | 50 (29-66) | 24-192 |
| C- reactive protein, mg/dL | 2.00 (0.85-5.03) | 1.87 (0.62-3.11) | 1.04 (0.45-4.95) | 2.77 (1.28-4.02) | 2.06 (1.17-5.59) | 0.1-1 |
| Procalcitonin, ng/mL | 0.068 (0.048-0.124) | 0.079 (0.073-0.084) | 0.048 (0.048-0.068) | 0.062 (0.058-0.131) | 0.081 (0.048-0.124) | 0-0.5 |
| Ferritin, ng/mL | 269.1 (179.4-955.6) | 128.1 (36.3-219.8) | 735.4 (185.4-1170.4) | 288.3 (221.5-682.7) | 269.1 (174.3-1057.9) | 30-400 |
| Troponine T, pg/mL | 15.18 (8.98-33.22) | 71.14 (50.87-91.40) | 6.37 (5.13-8.75) | 24.53 (11.77-46.18) | 14.77 (10.76-27.04) | <14 |

Fifty patients (96.2%) were treated with LMWH. Thirty-two patients (61.5%) received subcutaneous heparin at prophylactic doses, and 20 patients (38.5%) therapeutic doses. In the infected group, the use of heparin at prophylactic doses was more common than in the non-infected group (p=0.01). The overall median duration of treatment with LMWH was 11 days (IQR: 6.5 to 17.0).

Overall patient mortality was 1.9%; one patient died (6.3%) in the infected group, and none in the non-infected group (p=0.31).

## Discussion

We present our case series of patients with and without SARS-CoV-2 infection, who were studied by CTPA on suspicion of pulmonary thromboembolism (PE).

There have been several groups that have published their results studying SARS-CoV-2 infected patients with a suspicion of pulmonary embolus. Leonard-Lorant *et* al observed positive CTPA angiography for PE in thirty-two of 106 (30% [95%CI: 22-40%])^11^, with higher D-dimer levels than those without pulmonary embolus. They also observed that D-dimer greater than 2660 μg/L had a sensitivity of 32/32 (100%, 95%CI: 88-100) and a specificity of 49/74 (67%, 95% CI: 52-79) for pulmonary embolism on CTPA. Stoneham *et al* diagnosed 21 out of 274 (7.7%) patients with VTE, 16 with PE (5.83%)^12^. They found significance differences in levels of D-dimer and white cell count but not in age, gender, or presence of comorbidities between PE patients and control group. Klok *et al* detected 25 symptomatic VTE events in 184 adult patients admitted in three ICU across the Netherlands^13^. A similar study of 150 critical patients from four ICUs in France, found 16.7% of patients with PE despite the anticoagulant therapy^14^. In the study of Middeldorp *et al* 39 patients (19.6%) were diagnosed with VTE, despite routine thrombosis prophylaxis^15^. Lodigiani *et al* detected PE in 10 (33%) of CTPA performed^16^. Grillet *et al* studied one hundred patients with severe respiratory COVID disease and/or comorbidities by CTPA^17^. Twenty-three (23%, [95% CI: 15-33%]) patients had acute PE. Compared with patients without PE, these patients were admitted in ICU and required mechanical ventilation more frequently.

We have detected PE in 28.6% of infected patients who were studied with CTPA. All these studies revealed the importance of early onset of anticoagulant therapy. In fact, chronic anticoagulation therapy at admission has been associated with a lower risk of the composite outcome (Hazard Ratio [HR] 0.29, 95%CI: 0.091–0.92), and patients diagnosed with thrombotic complications are at higher risk of all-cause death (HR 5.4; 95%CI: 2.4–12)^18^. Some authors have highlighted the possibility that some individuals infected with COVID-19 may be refractory to treatment with low molecular weight heparin, and it has been studied the use of tissue plasminogen activator in these cases^19^.

The main clinical symptoms of COVID-19 patients in the study of Li *et al*, similar as our results, were fever (88.5%), cough (68.6%), myalgia or fatigue (35.8%), expectoration (28.2%), and dyspnea (21.9%)^20^.

We have not seen differences in mortality between both groups. However, patients with COVID-19 infection, with a high risk of venous thromboembolism, had poorer outcomes than patients with a low risk, suggesting that these patients might require increased attention in case of rapid deterioration^21^. Global mortality rates of SARS-CoV-2 infection has been established in 5.7%^22^, while our overall mortality was 1.9%.

The statistically significant differences detected in infected and non-infected PE patients in total CK and troponin T levels should lead us to consider the presence of myocardial damage caused by SARS-CoV-2, which is not found in uninfected PE patients.

Based on our data and experience, LMWH therapy should be considered, with at least prophylactic dose, in all patients with COVID-19 pneumonia for a longer period than required for clinical resolution of the disease, considering the high risk of thromboembolic complications of SARS-CoV-2 infection. Additionally, CTPA is the cornerstone in both the diagnostic workup and follow-up of SARS-CoV-2 infection.

The main limitations of our study are the small sample size, and the lack of generalized study with CTPA in all the admitted COVID patients at hospital. It is possible that our incidence of PE, as in other studies, is probably highly underestimated due to the paucity of CTPA performed for PE during hospitalization, and most of the studies included only critical patients in Intensive Care Unit (ICU). In fact, the risk for VTE in COVID-19 patients is high, particularly in ICU patients. We therefore cannot establish a realistic prevalence for PE in all our COVID patients. Another limitation is the possible bias of patient selection by incorporating only the most severe or expected diagnosis of PE results, but this has been attempted to control by consecutive incorporation of all CTPA performed.

A considerable incidence of PE diagnosed by CTPA in SARS-CoV-2 infected patients in our hospital has been observed, despite thrombo-prophylaxis. The infected patients with PE should be examined for assessing myocardial damage.

## Data Availability

Availability of all data and materials
Data and material cannot be showed.

## Ethical Approval

The local ethics committee approved this study (approved May 25, 2020) and waived the need of informed consent. Personal data were dissociated and pseudoanonymized in the database for further statistical analysis by an independent expert.

## Authors Contributions

DE had full access to all the data in the study and takes responsibility for the integrity of the data and the accuracy of the data analysis. FJCH and LA contributed substantially to study design, data collection and data analysis. All authors helped with study interpretation, writing, and editing of the manuscript.

## Availability of data and materials

Data and material cannot be showed.

## Acknowledgement

This paper is part of thesis doctoral of Laura Álvarez in University of Granada (Spain), Doctoral Pharmacy Program at Pharmacy Department.

A sincere thank you to Juan Pablo Fernandez for his diligent proofreading of this paper.

